# A retrospective case-control study for Clinical Validation of mutated ZNF208 as a novel biomarker of fatal blast crisis in Chronic Myeloid Leukemia

**DOI:** 10.1101/2024.03.09.24304026

**Authors:** Nawaf Alanazi, Abdulaziz Siyal, Salman Basit, Masood Shammas, Aamer Aleem, Amer Mahmood, Sarah Al-Mukhaylid, Zafar Iqbal

## Abstract

The hallmark of Chronic Myeloid Leukemia (CML) is Philadelphia chromosome t(9:22), which leads to formation of BCR-ABL1 fusion oncogene. BCR-ABL1 induces genetic instability, causing the progression of chronic myeloid leukemia (CML) from the manageable Chronic Phase (CP-CML) to the accelerated phase (AP-CML) and ultimately to the lethal blast crisis (BC-CML). The precise mechanism responsible for CML progression are not well comprehended, and there is a lack of specific molecular biomarkers for advanced phase CML. Mutations in transcription factors (TFs) have a significant role in cancer initiation, relapses, invasion, metastasis, and resistance to anti-cancer drugs. Recently, our group reported association of a novel transcription factor, ZNF208, with CML progression and there was a dire need for clinical validation of this novel biomarker. Therefore, the aim of this study was to clinically validate mutated ZNF208 as a novel biomarker for CML progression in a larger cohort of AP- and BC-CML patients using control-case studies.

A total of 73 CML patients (N=73) from King Saud University Medical City Riyadh and King Abdulaziz National Guard Hospital, Al-Ahsa, Saudi Arabia were enrolled in the study (2020-2023), with the experimental group (cases) consisting of patients AP-CML (n=20) and BC-CML (n=12). The controls consisted of age/sex matched CP-CML (n=41). The study was approved by Research Ethics Committees of participating institutes and all patients provided informed consent for the study. Clinical evaluations for patients were conducted according to the guidelines established by the European LeukemiaNet in 2020. Targeted resequencing of ZNF 208 was employed using Illumina NextSeq500 instrument (Illumina, San Diego, CA, USA) and mutations confirmed using Sanger sequencing.

Both next generation sequencing as well as Sanger sequencing identified a novel missense mutation (c.64G>A) in novel ZNF208. in 56 (93.3) and12 (100) CP-, AP- and BC-CML patients respectively, while in none (0%) of CP-CML patients or healthy controls from genomic databases (p=0.0001). Therefore, our studies show that ZNF208 mutation (c.64G>A) is novel and very specific biomarker for AP-and BC-CML patients. ZNF208 and other such proteins may cause carcinogenesis by interacting with KAP-1 repressor to silence many target genes and thus may prove to be novel drug targets as well. Therefore, we recommend carrying out prospective clinical trials for further clinical validation of this biomarker for its utilization in clinical decision, investigating its precise role in cancer pathogenesis and investigate its potential for novel drug target in advanced phase CML patients.

**Simple Summary:** Chronic Myeloid Leukemia (CML) is a type of blood cancer caused by the BCR-ABL1 fusion oncogene, leading to genetic instability and other genetic changes. This results in advancement from a manageable Chronic Phase (CP) to an accelerated phase (AP) and finally a lethal blast crisis phase (BC). The mechanism of CML development is not well known, and there is a dearth of dependable shared molecular indicators. Transcription factors (TFs) are a class of molecules that, when altered, significantly contribute to the development and advancement of cancer, including relapses, invasion, metastasis, and resistance to anti-cancer drugs. Recently, ZNF208 was has been reported to be novel transcription factor gene associated with BC-CML. Here, we carried out clinical validation of ZNF208 as a novel biomarker of CML progression using targeted resequencing. ZNF208 mutation (c.64G>A) was detected in 0 (0%), 56 (93.3) and12 (100) CP-, AP- and BC-CML patients respectively (p=0.0001) demonstrating its very high specificity for AP- and BC-CML. This shows that ZNF208 mutation (c.64G>A) is a very specific biomarker for CML progression. We recommend prospective clinical trials for further clinical validation of this novel biomarker for CML progression.

## 1. Introduction

Chronic myeloid leukemia (CML) is a type of blood cancer that affects the stem cells responsible for producing blood cells. It is characterized by the excessive generation of non-functional granulocytes in the bone marrow [1]. CML is characterized by the existence of the Philadelphia chromosome t(9:22), which is a reciprocal translocation between the Abelson murine leukemia (ABL) gene on chromosome 9 and the breakpoint cluster region (BCR) gene on chromosome 22 [2]. The fusion leads to the constant expression of tyrosine kinase BCR-ABL on hematopoietic stem cells (HSCs), causing them to convert into leukemic stem cells (LSCs) and facilitating uncontrolled development and replication [3]. An inherent characteristic of chronic myeloid leukemia (CML) is the presence of genomic instability induced by BCR-ABL itself. This instability results in the emergence of more mutations in both BCR-ABL and other genes as the disease advances [4].

The milestone discovery of BCR-ABL tyrosine kinase inhibitors (TKIs), specifically imatinib, has been a significant advancement in the treatment of chronic myeloid leukemia (CML), making overall survival of CML patients equaling general public, at least in technologically advanced countries [1]. This breakthrough has resulted in reduction in the rate of progression of CML patients from the chronic phase to an accelerated phase (AP-CML) and ultimately to a fatal blast crisis (BC-CML) [5]. Regrettably, certain individuals experience resistance to TKIs, resulting in therapy failure and progression to BC-CML [6]. However, the exact mechanisms responsible for the progression of CML are not fully understood, and there is a scarcity of reliable molecular markers for progression of CML to accelerated and blast crisis phases [3]. It makes early detection of CML patients at risk of disease progression, specifically to blast crisis, as one of biggest issues in 21st century cancer medicine [5, 6].

Transcription factors (TFs) have been found to be responsible for initiation onset, invasion, drug-resistance, metastasis and progression, and hence morbidities and mortalty of a many types of cancers [7].Transcription factors (TFs) are proteins that selectively attach to particular DNA sequences, facilitating the transfer of genetic information from DNA to messenger RNA (mRNA) and thus function as regulators of gene expression [8]. The activity of transcription factors (TFs) can be modified in cancer through many direct and indirect mechanisms including chromosomal translocations, gene amplification, deletion, point mutations, altered gene expression, noncoding RNAs, DNA methylation and histone structure through epigenetic factors [9]. Although TFs are recognized to have a significant role in the development of acute leukemias, their specific contribution to the progression of CML is still largely understood.

Our laboratory recently showed a novel transcription factor ZNF208 with a missense mutation (c.64G>A) associated with the progression of chronic myeloid leukemia (CML) [10]. Keeping in view that no reliable molecular biomarker is currently in clinical practice to early detect the CML patients at risk of developing fatal blast crisis and their timely therapeutic intervention in order to stop or delay their acute transformation [3, 5, 6], there was an urgent need to clinically validate this important finding in a broader population of patients with advanced-phase (AP) and blast crisis (BC) CML, using control-case studies [10]. Hence, the objective of this investigation was to clinically authenticate mutant ZNF208 as a new biomarker for the progression of chronic myeloid leukemia (CML).

## 2. Materials and Methods

This study focused on recruiting and selecting patients with Chronic Myeloid Leukemia (CML) who were enrolled at King Saud University Medical City Riyadh and King Abdulaziz National Guard Hospital Al-Ahsa in Saudi Arabia (N=73). The study period spanned from January 2020 to July 2023. The study comprised a total of 41 patients diagnosed with chronic phase chronic myeloid leukemia (CP-CML) as control group. The experimental group consisted of 20 patients with Accelerated Phase (AP-CML) and 12 patients with Blast crisis CML (BC-CML). Initially, all CP-CML patients received imatinib mesylate (IM) as their first-line treatment. Patients who did not respond well to IM were subsequently administered 2nd and third line tyrosine kinase inhibitors (TKIs)[12].

The criteria for treatment responses were adopted from the European LeukemiaNet (ELN) recommendations 2020 [11]. The definitions of clinical stages of chronic myeloid leukemia were adopted as in previous ELN guidelines [12, 13]. Nevertheless, the presence of resistance to two tyrosine kinase inhibitors (TKIs), identification of a mutation in the kinase domain (KD) of the BCR-ABL1 gene, or the development of other chromosome abnormalities in Philadelphia chromosome positive (Ph+) cells (ACA) give rise to worries over the advancement of the disease [11]. Evaluation standards for measuring the effectiveness of treatment in individuals with chronic myeloid leukemia related to hematological responses and cytogenetic responses were based on previous ELN guidelines [12, 13] and studies carried out previously [14-16]. Nevertheless, molecular response definitions were adopted from ELN 2020 guidelines [12].

Overall survival referred to the duration of time from the initiation of IM treatment to either the date of the patient’s death or the most recent follow-up [17]. Progression-free survival (PFS) was evaluated from the initiation of IM treatment until the development of chronic myeloid leukemia (CML) into accelerated phase (AP) or blast crisis (BC), or until death occurred. On the last follow-up date, every patient who had experienced a sustained event up until the last day of the study was excluded from the data. The survival status of patients who were not present during the previous follow-up was confirmed by reaching out to them using their recorded personal information. The Kaplan-Meier Method was selected for the purpose of conducting survival analysis [18]. Standard terminologies (version 4.03) were used for the classification of hemolytic adverse effects [12, 19].

All procedures used in this investigation were approved King Abdullah International Medical Research Center (KAIMRC) and King Saud University Medical City, Riyadh, Saudi Arabia and ethical approval from these centres was obtained per their regulatuions. Each patient who was enrolled in the trial provided signed informed consent to take part. All of the Declaration of Helsinki’s codes were followed by the consent form [20].

Ten milliliters of peripheral blood were drawn for this investigation and placed in EDTA tubes. (BD Vacutainer Systems, located in Franklin Lakes, New Jersey.) The QIAamp DNA Blood Mini Kit (QIAGEN) was utilized for DNA extraction, and the NanoDrop Spectrophotometer (NanoDrop Technologies, Inc., USA) was employed for DNA quantification. After dilution, aliquots of 70–80 ng/μl were created in order to detect mutations using Whole Exome Sequencing (WES). To perform Sanger Sequencing, the residual DNA has to be diluted to 40 ng/μl. The DNA samples were kept in a refrigerator at -80°C [21-22].

Next Generation Sequencing (NGS) was used to do targeted resequencing of the ZNF208 gene.

In order to depict each stage of the disease (Chronic Phase and Accelerated Phase), carefully selected and thoroughly analyzed samples of CML were utilized for NGS. The Illumina® DNA Prep with Enrichment, (S) Augmentation kit (Cat. # 20025523) was used to enrich the target DNA for ZNF208 [22, 23]. The initial stage of Next-Generation Sequencing (NGS) involved the process of DNA fragmentation, which was then followed by tagmentation. Subsequently, the DNA fragments that had been tagged were amplified and subsequently purified using magnetic beads. Subsequently, Oligos were employed to capture specific regions of interest. The amplified libraries were enriched using PCR and their quantity was determined using a Qubit fluorometer. The library size distribution was measured using an Agilent Bioanalyzer. The Illumina NextSeq500 instrument was used to perform cluster generation and exon sequencing. The quantified DNA libraries were loaded onto the flow cell for this purpose [22, 23].

### Analysis of Next Generation Sequencing (NGS) Data

The BCL2FASTQ software was used to convert the output files, specifically the BCL files, into FASTQ files. The BWA Aligner was used to align the FASTQ data to the human genome, utilizing the BWA-MEM algorithm. The Genome analysis tool kit (GATK) was used to identify variants. The genomic variations were annotated and filtered using Illumina Variant Studio [22,24].

Primary Analysis: The ZNF208 gene was examined in all patients with AP-CML to identify common biomarkers associated with the progression of chronic myeloid leukemia (CML). Filtering techniques that utilized the identification of uncommon genetic variations and the exclusion of intron and synonymous variations were employed to change the excel file that presents Next-Generation Sequencing (NGS) data. In addition, any variations that had a known prediction of being either benign (B) or tolerant (T) were eliminated. According to Carson (2014), certain variations were labeled as B if they had a frequency of 70% or higher for B, while other variants were categorized as T if their frequency for T was 70% or higher [25]. Any variants with a population frequency greater than 0.005 in the dbSNP and Exome Sequencing Project (ESP) database were also excluded. Therefore, the process of identifying variants was restricted to only those with significant protein impacts and splice variants. In addition, the data was subjected to additional analysis in order to examine new genetic variants that are found in patients with advanced phase chronic myeloid leukemia (AP-CML & BC-CML), but not in patients with chronic phase chronic myeloid leukemia (CP-CML) or healthy individuals. This suggests that these mutations may have a role in the progression of the disease [26, 27].. The data generated by next-generation sequencing can be accessible from the National Center for Biotechnology Information (NCBI), where it was submitted.

### Sanger sequencing to validate mutations

Sanger sequencing was used to verify the variations identified by WGS. The University of California Santa Cruz’s genome database browser was used to retrieve the genomic primers for the variations found in the indicated genes (Table 2); primers themselves were from Applied Biosystems in California, USA. The process of amplifying Template DNA involved PCR. DNA sequencing reactions were performed using ABI PRISM Big Dye Terminator Cycle Sequencing Ready Reaction kits (Applied Biosystems, California, USA) [28]. The ABI Prism 3730 Genetic Analyzer (Applied Biosystems, California, USA) (http://genome.ucsc.edu/cgibin/hgGateway) was used to perform Sanger sequencing on both forward and reverse DNA templates [29, 30].

**Table 1.**
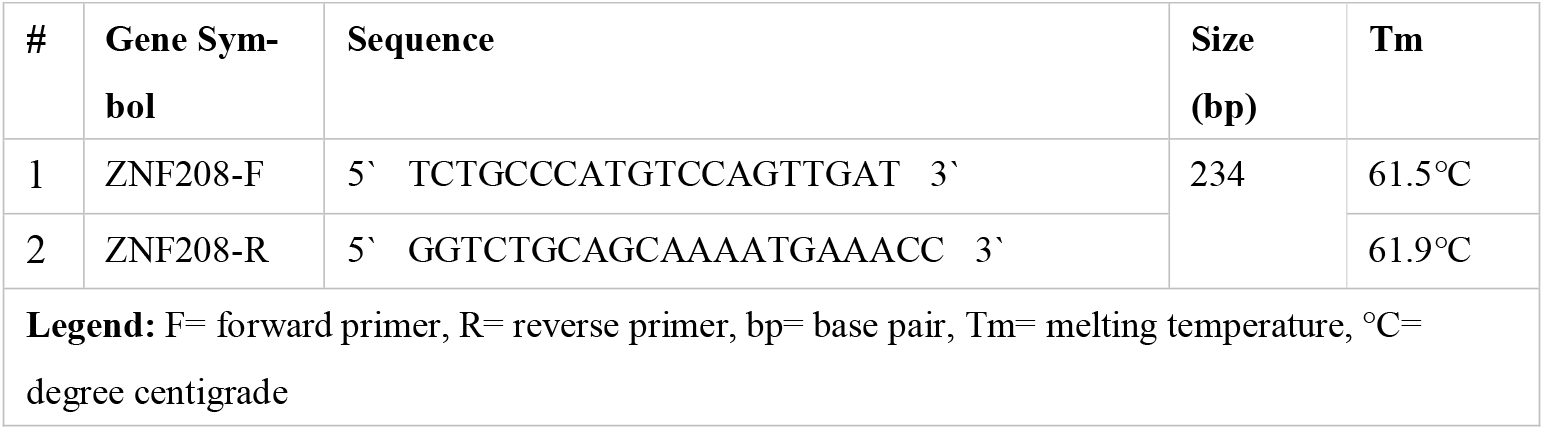
Common variants in Accelerated/Blast Phase-Chronic Myeloid Leukemia patients (AP, n=5, BC; n=7), primers designed to amplify the regions around the detected variants in FANCD2 and ZNF208 genes.

**Table 2.** Demographics and Laboratory Parameters In Our Study.

| Characteristics | # of Patients, % |
| --- | --- |
| Total (n=73) |  |
| Age, yrs |  |
| Mean (range) | 34.6 |
| Gender |  |
| Male | 44 (63%) |
| Female | 29(39.37% |
| Ratio: Male: Female | 1.6:1 |
| Hemoglobin (g/dL), Mean | 10.1 |
| WBC count ($\times 10^9/L$), Mean | 313.7 |
| Platelets ($\times 10^9/L$) (n =108), Mean | 400.2 |

### Analysis of patient clinical data using statistical methods

The normalcy test provided evidence for percentages, absolute numbers, and categorical data. For continuous data, the same procedure was used in addition to a suitable measure of variance. We used Fisher’s exact test or Chi-Square test to compare two means of categorical data. The Mann-Whitney U test or a two-sample independent test were utilized for continuous data. Furthermore, ANOVA or Kruskal-Wallis tests were performed to ascertain the variance for groups of ≥3. The survival outcome was measured using Kaplan-Meier survival analysis curves [18]. The group comparison was conducted using the log-rank test. Version 9.4 of the SAS/STAT software (SAS Institute Inc., Cary, NC, USA) was used for data analysis. For statistical computation, the R foundation was used (Vienna, Austria) [31]. The Sokal risk score, Eutos risk score, and Euro risk score were calculated [11, 32–35].

## Results

### 3.1 Demographics

A total of 73 patients with CML were enrolled in this study. The mean age was 34.6 years, and the male-to-female ratio was 1.6:1, with 60% males and 39.8% females. The mean hemoglobin, WBC count, and platelets were 10.1, 317.9, and 400.2, respectively (Table 2). Overall, males were more commonly affected by CML.

The mean age of CP-, AP-, and BC-CML patients was 33.5, 35.6, and 38.1 years, respectively. Additionally, all CML phases were dominated by males, and the male-to-female ratio was 1.6:1, 1.9:1 and 2:1, in CP-, AP- and BC-CML patients, respectively, with more trend in CML progression in male patients than female though differences were not statistically different (Table 3). CP, AP and BC-CML patients showed statistically significant differences with respect to leukocyte count of 50 × 109/L or higher (p=0.04), splenomegaly of spleen size 5cm or more, hepatomegaly (p=0.006), mortality (p=0.001) and survival (p=0.001). Nevertheless, as none of these variables was helpful in early prediction of CML progression decisively, molecular genetic analysis of already reported ZNF208 gene was carried out to find out the genetic basis of CML progression and for validation of mutated ZNF208 as new biomarker of CML progression.

**Table 3:**
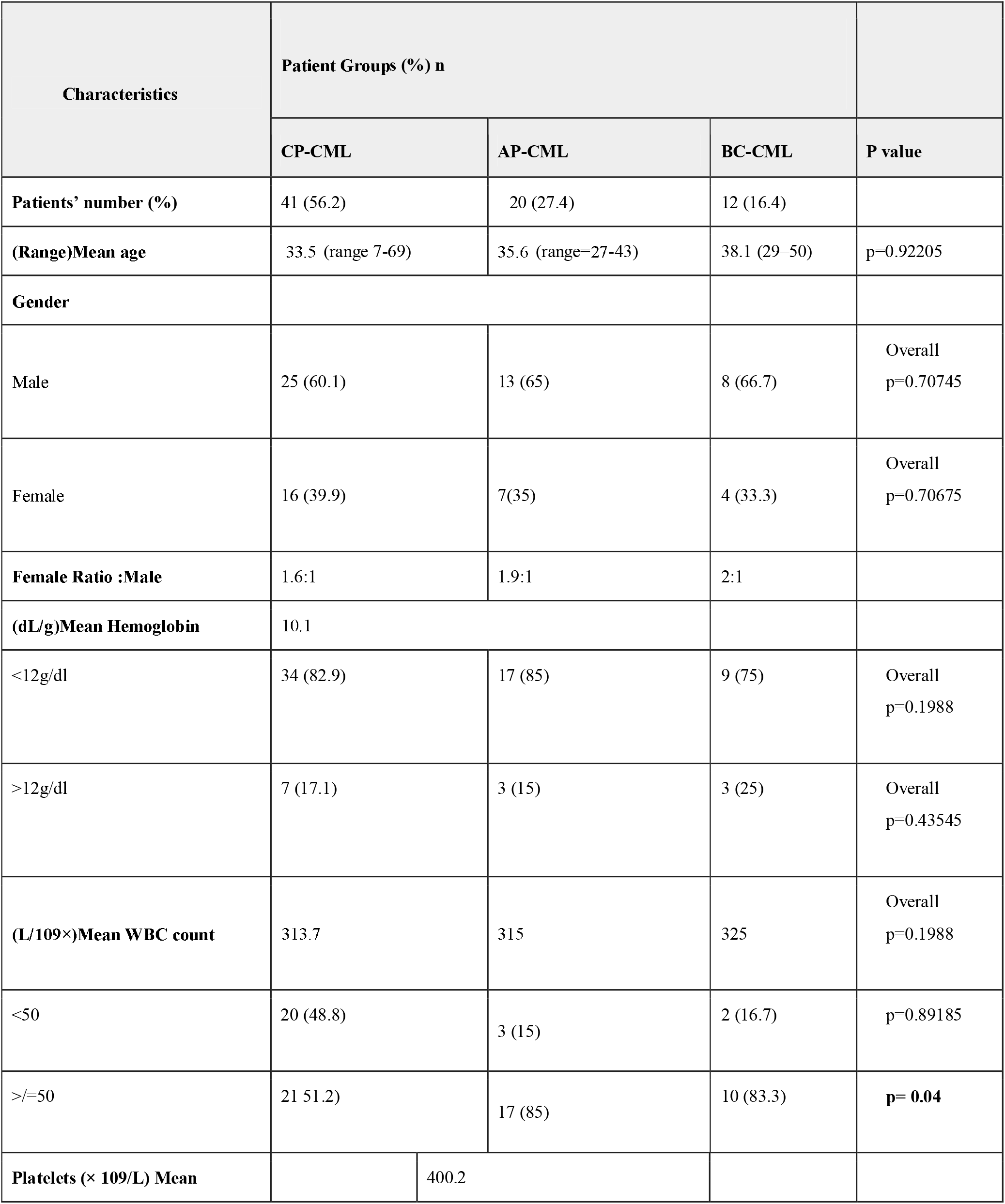
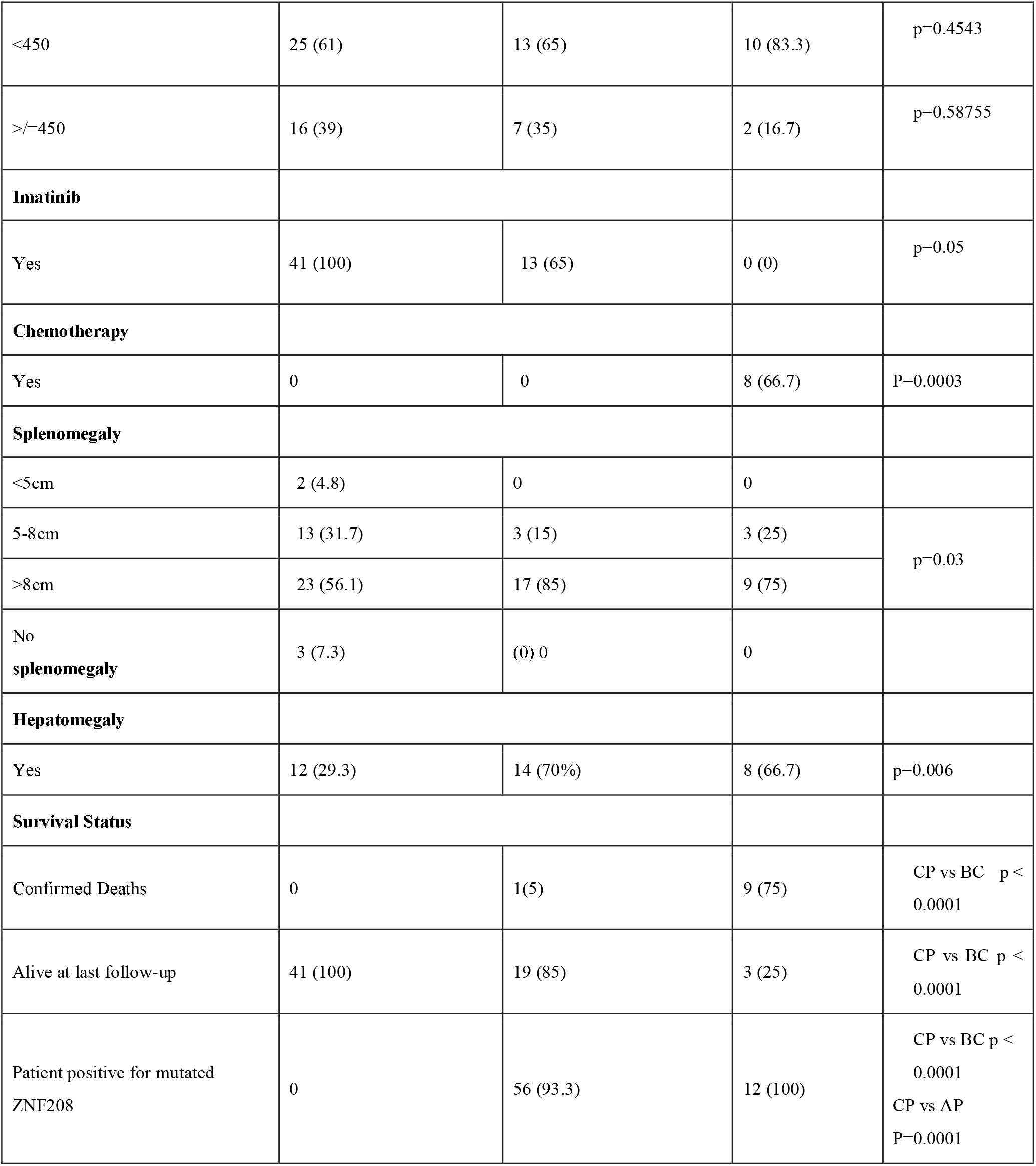
A comparison between CP- and AP-CML patients in this study in regard to their demographic and laboratory characteristics.

### 3.2 Clinical validation of ZNf208 by Targeted resequencing

*Targeted resequencing* employed to validate ZNF208 as novel biomarker of CML progression in cases (AP- and BC-CML patients) and controls (CP-CML patients) detected ZNF208 64G>A) mutaions in 0%, 93.3% and 100% of CP-, AP- nad BC-CML patients respectively. The results were confirmed by Sanger sequencing. It shows that ZNF208 is a very useful marker of CML progression with specifcity of 93% to 100%.

The mutation detected in ZNF208 is new point mutation (c.64G>Ain which Guanine was replaced by Adenine at the at condon 22 at position 64 downstream of ZNF208 promoter. This change resulted in Asparagine instead of Aspartate at protein level.

**Figure 2:**
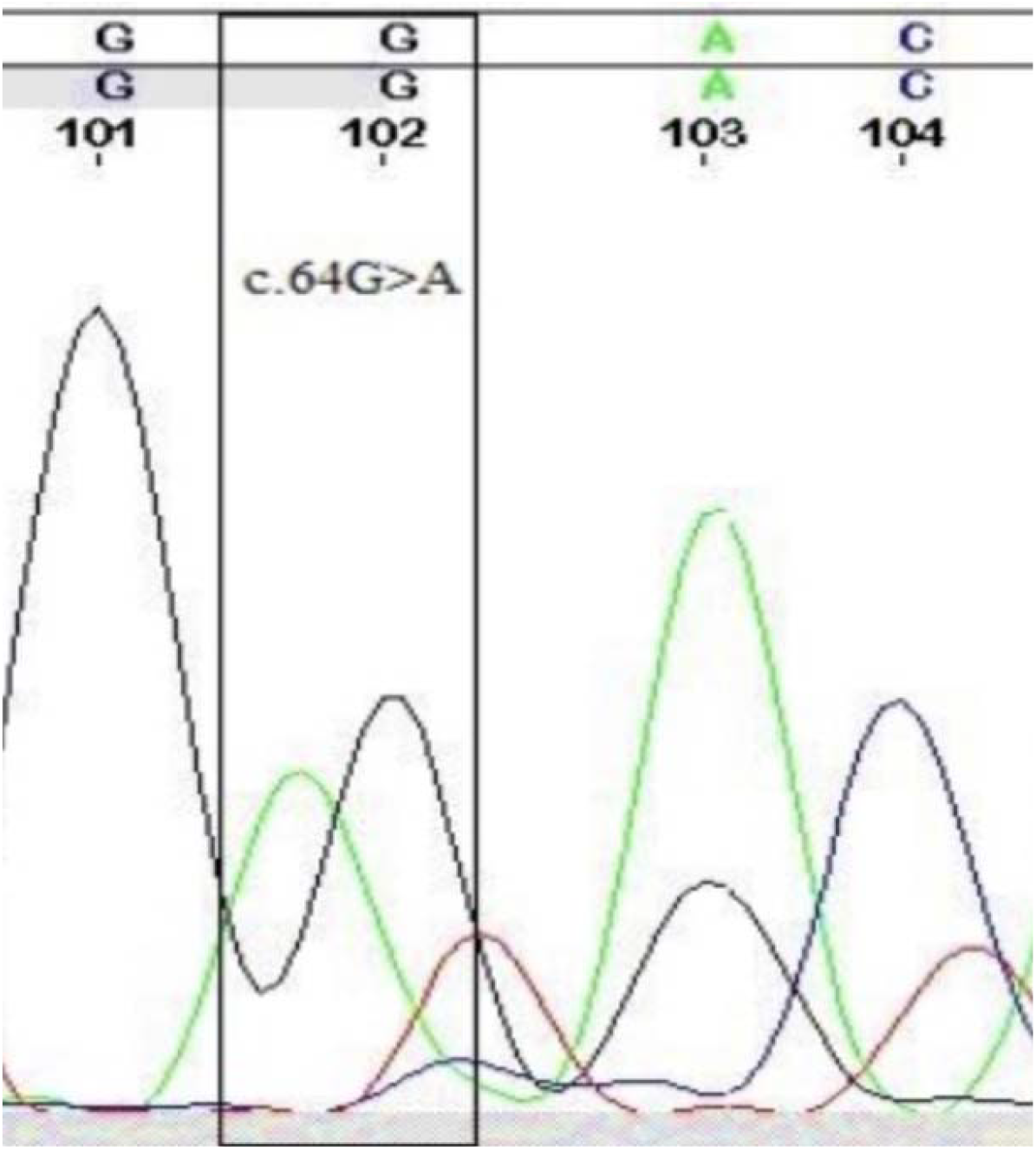
Validation of missense mutation ZNF208 (c.64G>A) by Sanger sequencing.

Thus, it is concluded that ZNF208, a member of the zinc-finger binding transcription factor family, is mutated in all BC-CML and 93% AP-CML patients, with an impact on change in znf208 protein sequence, thus suggesting it to be a “likely to be pathogenic” mutation and a very important and novel molecular biomarker for CML progression that can help in early identification of patients at risk of ftransformation to fatal BC-CML and thus prividing the oncologists opprtunity to therapeutically intervence such high=risk CML patients using a varierty of newly developed BCR-ABL inhibitors The study shoul be furthr extended to unravel the role of ZNF208 in CML pathogenesis and prospective clinical trials should be carried out to further evaluate the role of ZNF208 G64A mutation and other such mutations of this important gene as a novel biomarker and primising drug target for AP- and BC-CML.

## 4. Discussion

This study included a total of 73 as case-control subjects that were from different phases of chronic myeloid leukemia (CML). The case-control based clinical validation of potential disease biomarker is a gold standard in discovery and clinical validation of novel biomarkers [36, 37]. We used NGS-based targeted resequencing and Sanger sequencing for clinical validation of ZNF G64A mutation as AP-/BC-CML. Targeted resequencing and Sanger sequencing along with other similar techniques are routinely being used for clinical validation of molecular biomarkers [38-45]. Moreover, incorporation of next-generation sequencing, and other high-throughput techniques have refined the discovery and validation of diagnostic and prognostic biomarkers and their incorporation into precision medicine [43-51].

Our findings revealed that the ZNF208 gene remained unaltered in chronic phase CML patients as well as in healthy individuals. However, it was shown to be mutated in all patients in the advanced phase. The relationship between CML advancement and the development of a lethal blast crisis can be attributed to ZNF208, a member of the biologically significant group of genes known as Zinc Finger Proteins. The human genome contains a large number of zinc finger (ZNF) genes, with around 500-600 members [50]. These proteins have a role in controlling genes and the process of development, and they have remained mostly the same throughout evolution [51].

Research has indicated that the aberrant synthesis of many ZNF proteins [52] has an impact on cancer development through several mechanisms. ZNF309 has been detected in various types of cancer, including colorectal cancer, multiple myeloma, prostate cancer, cervical cancer, colon cancer, ovarian cancer, and neuroblastoma [53]. An relationship was discovered between elevated gene expression of ZNF-281 and many cancers including pancreatic, colon, breast, neuroblastoma, and ovarian malignancies. The findings indicated that there was an overexpression of the ZNF-281 protein, which was connected with the development of the disease [53]. However, there was a reduction in the expression of ZNF-281 observed in relation to the development of cancer. Specifically, while excessive expression of ZNF-281 may be associated with a negative prognosis, it actually suppresses the development of cancer in glioma and non-small cell lung cancer, but in a contrasting manner [52]. ZNF proteins have recently been identified as protectors of genomic integrity. Mutations in ZNF genes can cause genomic instability through well-known mechanisms [54].

As previously mentioned, we identified a missense mutation in the ZNF208 gene that is specifically linked to patients with advanced phase and blast crisis chronic myeloid leukemia (CML). This mutation results in the substitution of Aspartate with Asparagine in the ZNF208 protein. The ZNF208 gene is situated on Chromosome 19 in the p12 region [55]. It interacts with DNA and regulates the process of gene transcription [50, 56]. Furthermore, additional research has demonstrated a correlation between ZNF208 and various other medical conditions. A Genome-Wide Association Study (GWAS) identified a correlation between telomere length and the ZNF208 gene [57]. A study conducted in China indicates a correlation between Single nucleotide polymorphisms (SNPs) in ZNF208 and susceptibility to Hepatitis B virus (HBV). The study included of 300 healthy volunteers and 242 patients who tested positive for HBV. The results suggest that ZNF208 variations may have a substantial impact on the progression of HBV. The study found that ZNF208 polymorphisms significantly increased susceptibility to HBV infection (P = 0.008) [50]. Furthermore, a scientific investigation has asserted that mutations in the ZNF208 gene are associated with an inclination towards developing esophageal cancer [58]. According to another study, the likelihood of developing laryngeal cancer increases when there are particular variations in the ZNF208 gene, specifically the rs8103163 A and rs7248488 A alleles [59]. A separate Chinese study investigated the correlation between Chronic obstructive pulmonary disease (COPD) and genetic variations in ZNF208. The study recruited 270 people with chronic obstructive pulmonary disease (COPD) and 288 individuals who were in good health. The study indicated that ZNF208 variations had an impact on telomere length in individuals with COPD [60]. Recently, it has been discovered that cirZNF208 has a function in the development of cervical cancer and has an impact on its prognosis. This demonstrates the significance of ZNF208 in the field of biology, particularly in relation to health and illness. It provides valuable insight into the vital function that ZNF208 plays in the progression of CML.

Mutations in the ZNF208 gene have the potential to serve as a new biomarker for tracking the progression of chronic myeloid leukemia (CML). Additionally, these mutations can aid in tailoring tailored treatment for patients with advanced phase CML and other types of cancer. ZNF 432 has recently been discovered to function as an effector of HR-based DNA repair. This finding suggests that PARylation may play a regulatory role in this mechanism [54]. PARYlation is known to co-ordinate the recruitment of crucial proteins involved in the DNA damage response and guide the DNA repair pathways. The expression of ZNF432 was reduced in cases of resistance to the PARP inhibitor Olaparib. Additionally, altering the expression of ZNF432 in ovarian cancer cells was observed to increase their susceptibility to PARP inhibitors such as Olaparib [54]. This study demonstrates that a growing collection of zinc finger proteins (ZFPs) have been discovered to affect the stability of the genome. These ZFPs have the potential to be used in the development of a predictive gene signature, which could have prognostic value in relation to ovarian cancer and its response to PARP inhibitors. The sources cited for this information are [54, 62, 63-66].

Some novel investigations have unraveled the mechanism of ZNF-mediated repression of transcription in genome. The proteins coded by ZNF genes have a Canonical Kruppel-associated box (KRAB) domain that has affinity for KRAB-associated protein-1 (KAP-1) or Trim28 [67]. KRAB-ZFP identifies a specific region in the genome that is intended for suppression. The engagement with dimeric Trim28 subsequently attracts the epigenetic apparatus to achieve strong transcriptional suppression [67-69]. As ZNF208 G64A mutation under our investigation also lies in KRAB domain of znf208 protein, it may have a role in KAP1-mediated repression of some other target proteins in the genome, leading to CML progression in our subset of CML patients and utilizing agonists of ZNFs and KP-1 can lead to development of novel anti-cancer drugs [69-70]. These studies show that by integrating multi-omics with artificial intelligence (AI), further studies can expedite the cancer drug development for treating the currently fatal phenotypes of cancers like blast-crisis CML [43-49. 71].. Therefore, we recommend further investigations into role of mutated ZNF208 in CML progression, its validation as biomarker in prospective clinical trials and validating role of this and other such transcription factor as potential drug target in leukemia, cancers and other deadly diseases.

Therefore, it can be concluded that ZNF208, which is a member of the zinc-finger binding transcription factor family, is mutated in all patients with BC-CML and in 93% of patients with AP-CML. This mutation has an effect on the sequence of the ZNF208 protein, which suggests that it is a “likely to be pathogenic” mutation. Furthermore, it is a very important and novel molecular biomarker for the progression of CML. It can assist in the early identification of patients who are at risk of ftransformation to fatal BC-CML, thereby providing oncologists with the opportunity to therapeutically intervene in such high-risk CML patients by utilizing a variety of newly developed BCR-ABL inhibitors. In order to unravel the role that ZNF208 plays in the pathogenesis of chronic myelogenous leukemia (CML), the study should be further extended. Additionally, prospective clinical trials should be carried out in order to further evaluate the role of the ZNF208 G64A mutation and other similar mutations of this important gene as a novel biomarker and primary drug target for acute myelogenous leukemia (AP- and BC-CML).

## 5. Conclusions

We concluded that in our patient population, CML progression occurred at a high rate. In advanced-phase CML patients, we discovered a novel transcription factor gene associated with CML progression using next-generation sequencing, pointing to a shared mechanism underlying CML’s abrupt transformation. Future research is necessary to confirm the functional genetic role of the gene in this study, and its evaluation as an early biomarker of CML disease progression. This will undoubtedly be useful to utilize this gene as a biomarker to determine subgroups of patients at risk of development of blast crisis and to clinically intervene in acute CML transformation to halt disease progression. To confirm our findings in a broader patient population and determine the precise function of this gene in the development of CML, more research is necessary. By using cutting-edge cellular and molecular biological techniques, the potential of ZNF208 as a novel therapeutic target for advanced-phase CML should also be investigated.

## Data Availability

All data produced in the present study are available upon reasonable request to the authors

## Author Contributions

Conceptualization, Nawaf Alanazi and Zafar Iqbal; Data curation, Salman Basit, Formal analysis, Zafar Iqbal; Investigation, Nawaf Alanazi,, Salman Basit, and Zafar Iqbal; Methodology, Sarah AlMukhaylid, i; Project administration, Zafar Iqbal; Resources, Nawaf Alanazi, Salman Basit and Zafar Iqbal; Software, Salman Basit, Masood Shammad, Amer Mahmood, and Zafar Iqbal; Supervision, Zafar Iqbal; Validation, Amer Mahmood; Visualization, Sarah AlMukhaylid; Writing – original draft, Aamer Aleem,, Sarah AlMukhaylid,; Writing – review & editing, Masood Shammad, Aamer Aleem,, Sarah AlMukhaylid, and Zafar Iqbal.

## Institutional Review Board Statement

The study was conducted in accordance with the Declaration of Helsinki, and was approved by King Abdullah International Medical Research Centre (KAIMRC), National Guard Health Affairs, Saudi Arabia, although no research funding was provided (project # RA17/002/A). This study was partially supported by the College of Medicine, Research Centre, Deanship of Scientific Research, King Saud University, Riyadh, Saudi Arabia. All authors have read and have consented to the acknowledgment.

## Informed Consent Statement

Informed consent was obtained from all subjects involved in the study.

## Data Availability Statement

Data generated from next-generation sequencing have been submitted to NCBI and can be accessed at https://www.ncbi.nlm.nih.gov/sra/PRJNA734750 (SRA accession number PRJNA734750), accessed on 18 Oct 2023.

## Acknowledgments

The study was approved by King Abdullah International Medical Research Centre (KAIMRC), National Guard Health Affairs, Saudi Arabia, although no research funding was provided (project # RA17/002/A). This study was partially supported by the College of Medicine, Research Centre, Deanship of Scientific Research, King Saud University, Riyadh, Saudi Arabia. All authors have read and have consented to the acknowledgment. This project was financed by the National Plan for Science, Technology, and Innovation (MAARIFAH), King Abdul-Aziz City for Science and Technology, Kingdom of Saudi Arabia, Grant Number 14-Med2817-02.

